# Release of infectious virus and cytokines in nasopharyngeal swabs from individuals infected with non-B.1.1.7 or B.1.1.7 SARS-CoV-2 variants

**DOI:** 10.1101/2021.05.20.21257393

**Authors:** Blandine Monel, Delphine Planas, Ludivine Grzelak, Nikaïa Smith, Nicolas Robillard, Isabelle Staropoli, Pedro Goncalves, Françoise Porrot, Florence Guivel-Benhassine, Nathalie Demory Guinet, Julien Rodary, Julien Puech, Victor Euzen, Laurent Bélec, Galdric Orvoen, Léa Nunes, Véronique Moulin, Jacques Fourgeaud, Maxime Wack, Sandrine Imbeaud, Pascal Campagne, Darragh Duffy, James P. Di Santo, Timothée Bruel, Hélène Péré, David Veyer, Olivier Schwartz

## Abstract

The mechanisms that allowed for the SARS-CoV-2 B.1.1.7 variant to rapidly outcompete pre-existing variants in many countries remain poorly characterized. Here, we analyzed viral release, anti-SARS-CoV-2 antibodies and cytokine production in a retrospective series of 427 RT–qPCR+ nasopharyngeal swabs collected in COVID-19 patients harbouring either non-B.1.1.7 or B.1.17 variants. We utilized a novel rapid assay, based on S-Fuse-T reporter cells, to quantify infectious SARS-CoV-2. With both non-B.1.1.7 and B.1.1.7 variants, viral titers were highly variable, ranging from 0 to >10^6^ infectious units, and correlated with viral RNA levels. Lateral flow antigenic rapid diagnostic tests (RDTs) were positive in 96% of the samples harbouring infectious virus. About 67 % of individuals carried detectable infectious virus within the first two days after onset of symptoms. This proportion decreased overtime, and viable virus was detected up to 14 days. Samples containing anti-SARS-CoV-2 IgG or IgA did not generally harbour infectious virus. The proportion of individuals displaying viable virus or being RDT-positive was not higher with B.1.1.7 than with non-B.1.1.7 variants. Ct values were slightly but not significantly lower with B.1.1.7. The variant was characterized by a fast decrease of infectivity overtime and a marked release of 17 cytokines (including IFN-β, IP-10, IL-10 and TRAIL). Our results highlight differences between non-B.1.1.7 and B.1.1.7 variants. B.1.1.7 is associated with modified viral decays and cytokine profiles at the nasopharyngeal mucosae during symptomatic infection.

## Introduction

SARS-CoV-2 variants have supplanted pre-existing viral strains in many countries. The B.1.1.7 variant, initially detected in UK, displays a 43-90% higher reproduction number than pre-existing variants and is on track to become dominant worldwide ^1,2^. The reasons for this increased transmission are not fully deciphered. Proposed mechanistic hypotheses, based on epidemiological modelling, include higher viral loads and longer infectious period, rather than increased susceptibility in children or reduced protection from immunity to pre-existing variants ^1,2^. Little is known about the extent of virus shedding in individuals infected with B.1.1.7. A longitudinal study performed on a limited number of patients (7 individuals were tested) reported that the variant may cause longer infections with similar peak viral RNA concentration compared to non-B.1.1.7 SARS-CoV-2 ^3^. It has alternatively been observed that B.1.1.7 is associated with higher viral loads than non-B.1.1.7 viruses, as assessed by RT-qPCR ^4,5^. None of these studies examined infectivity of the specimen. Whether B.1.1.7’s increased transmissibility is linked to inherent viral properties such as the known increased affinity to ACE2, extended duration of shedding, higher viral peaks, or modifications of the local inflammatory state, remains to be mechanistically determined.

The extent and duration of shedding of infectious virus in nasopharyngeal swabs of COVID-19 patients are only partially characterized. Some studies have corelated RT-qPCR levels, or a positive lateral flow antigenic rapid diagnostic test (RDT), with viral outgrowth assays ^6-15^. Assessment of viral infectivity is classically performed using subclones or derivatives of the Vero cell line, which is naturally sensitive to infection and does not mount a type-I Interferon response ^6-12,16^. In these cells, the presence of replicating virus is generally detected by visualisation of a cytopathic effect after 2-10 days of culture, or by immunofluorescence staining or RT-qPCR measurement at an earlier time point. Clinical samples are often tested at low dilutions (pure to 1/10), precluding the calculation of an infectious titer. However, Bullard et al titrated a series of 90 swabs on Vero cells, and positive cultures were observed up to day 8 after symptom onset, with a median TCID50/mL of 1780 ^13^. With the same technique, Pickering et al reported titres up to 10^5^ PFU/mL on a series of 141 specimen ^14^.

Here, we explored virological and immunological characteristics of a retrospective series a 427 RT-qPCR positive nasopharyngeal swabs from individuals infected with non-B.1.1.7 and B.1.1.7 variants, collected at different days post onset of symptom (POS) and highlight potential differences between viral strains.

## Results

### Measuring virological and immunological parameters in nasopharyngeal swabs

We examined the content of nasopharyngeal swabs from RT-qPCR+ COVID-19-diagnosed individuals with four different assays (Fig. 1a). We assessed SARS-CoV-2 antigenic reactivity with a lateral flow antigen rapid diagnostic test (RDT) (from SD Biosensor Inc). We measured anti-SARS-CoV-2 IgG and IgA with the sensitive flow-cytometry based S-Flow assay ^17^. In a subset of 202 samples, we measured the levels of 48 cytokines, by either a bead-based multiplexed immunoassay system Luminex or the digital Elisa Simoa ^18^. We also titrated infectious virus with the S-Fuse assay ^19,20^. S-Fuse cells are U2OS cells stably expressing ACE2 and including the GFP–Split complementation system ^21^, producing GFP upon cell-cell fusion. The total area of syncytia, measured at 20h post infection with a high-content imager, correlates with the viral inoculum, indicating that the assay provides a quantitative assessment of viral infection ^20^. We improved the sensitivity of viral detection by stably expressing TMPRSS2, a surface serine protease known to enhance infection, in the cells, yielding S-Fuse-T cells. We first tested 12 RT-qPCR negative and 17 RT-qPCR positive nasopharyngeal swabs (Fig. 1b). The negative samples did not generate a GFP signal. Serial dilutions of RT-qPCR positive specimen demonstrated substantial heterogeneity in infectivity (from 0 to 10^5-6^ Infectious Units (IU)/ml). Limiting dilutions of 20 nasopharyngeal swabs demonstrated that S-Fuse-T cells were similarly sensitive to Vero E6 cells (assessed by the cytopathic effect generated at 5 days pi) to detect viral infectivity, with a strong correlation between the two assays (Fig. 1b). A representative experiment with two positive samples is depicted in Fig. 1c. Of note, viral release in the supernatant of S-Fuse-T infected cells reached only 10^3^ pfu/ml, probably because the cells rapidly die upon infection.

**Fig. 1.**
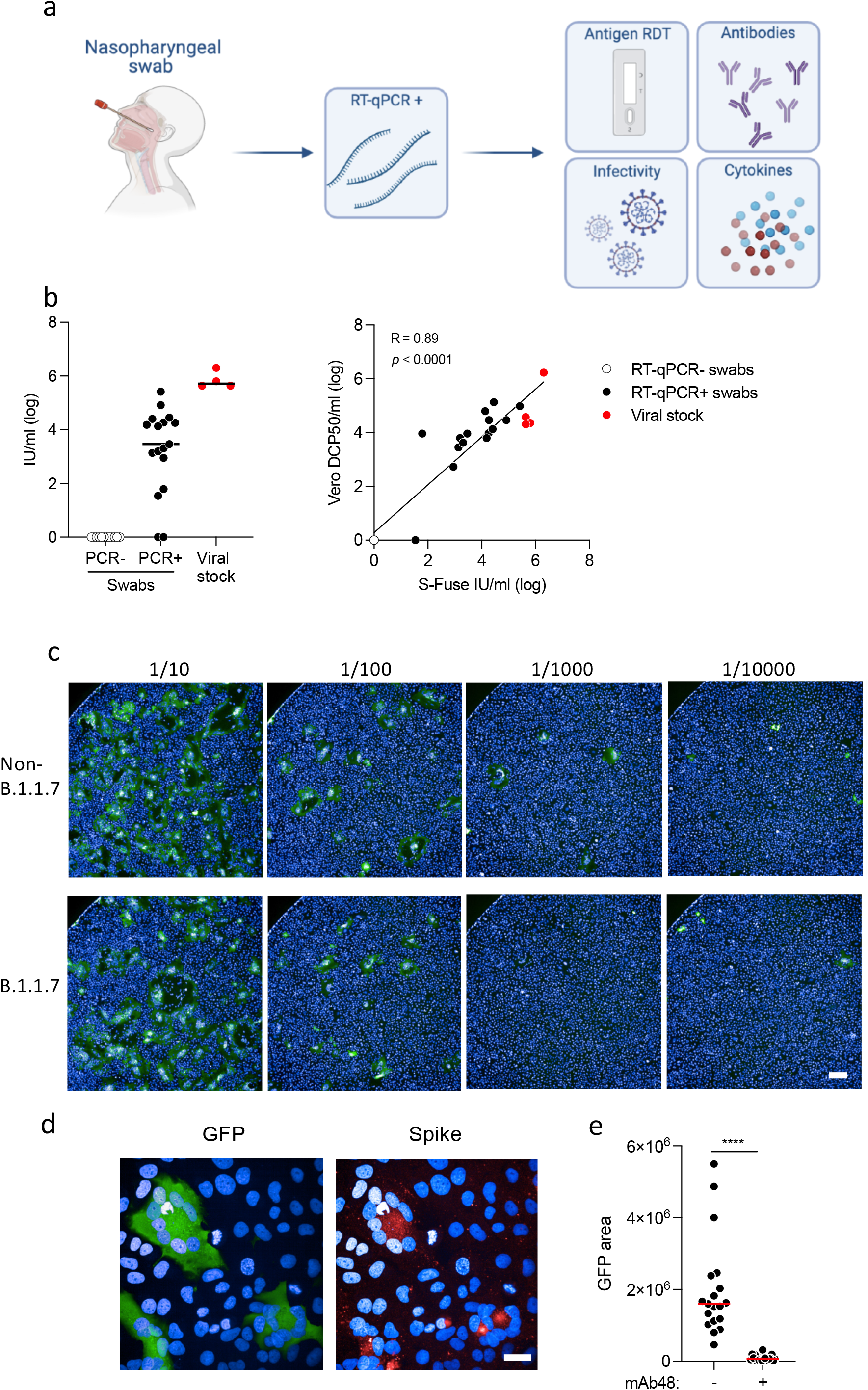
Detection of infectious virus, antibodies and cytokines in nasopharyngeal swabs. **a**. Study design. A retrospective series of 427 RT–qPCR+ nasopharyngeal swabs from COVID-19 patients, harboring non-B.1.1.7 or B.1.17 variants, was analyzed. Four tests were performed: a lateral flow antigen rapid diagnostic test (RDT), anti-SARS-CoV-2 IgG and IgA were measured with the flow-cytometry based S-Flow assay, infectious virus was titrated with the S-Fuse assay, and 46 cytokines were quantified in a subset of 202 swabs (70 non-B.1.1.7 and 132 B.1.1.7 samples) by Multiplex or Simoa assays. **b**. Titration of infectious SARS-CoV-2 in the swabs. Left panel: Titration was performed with S-Fuse-T cells, which become GFP+ after infection. S-Fuse-T cells were exposed to serially diluted swabs (10^−1^ to 10^−5^) or to purified viral stocks (red dots) as a control. 12 RT–qPCR-(white dots) and 17 RT–qPCR+ (black dots) samples were first analyzed. An infectious titer was calculated in Infectious Units (IU)/ml, after automatic scoring of the area of GFP+ cells at each dilution. Right panel: correlation between titers measured in Vero cells (in DCP50/ml) and S-Fuse-T cells (in IU/ml). **c**. Representative images of S-Fuse-T cells exposed to the indicated dilutions of nasopharyngeal swabs. Samples from one non-B.1.1.7-infected and one B.1.1.7-infected individuals are shown. Scale bar: 400 µm. **d**. GFP (green) and S (red) expression in S-Fuse-T cells exposed to one infectious swab analyzed by immunofluorescence. The Hoechst dye (blue) stains the nuclei. Scale bar: 40 µm. **e**. Neutralization of infectious virus by mAb48. Swabs (n=19) were preincubated 30 min at RT with mAb48 (1 µg/ml) and added to S-Fuse-T cells. Result from one representative experiment out of 3 is shown. A Wilcoxon paired t-test was performed **** p<0.0001.

We counterstained fixed plates with mab48, a human neutralizing monoclonal antibody targeting the SARS-CoV-2 Receptor Binding Domain (RBD) of the Spike, which does not cross-react with seasonal coronaviruses ^20^. The GFP+ cells also expressed the Spike (Fig. 1d), indicating that they have been productively infected with SARS-CoV-2 and not with another respiratory virus that may have triggered syncytia formation. Moreover, pre-incubation of the nasopharyngeal swabs with mab48 neutralized infection and inhibited the GFP+ signal, confirming that S-Fuse-T cells assayed infectious SARS-CoV-2 (Fig. 1e). Thus, S-Fuse-T cells provide a rapid system to detect infectious SARS-CoV-2 in nasopharyngeal swabs.

### Characteristics of COVID-19 individuals

We screened two series of nasopharyngeal swabs from RT-qPCR diagnosed COVID-19 individuals. Samples were collected at Hôpital Européen Georges Pompidou (Paris, France) for diagnostic purposes and further analyzed in accordance with local ethical standards. The biological (age and sex) and clinical (disease severity and days POS) parameters of the individuals are depicted table S1. The first series is composed of 200 samples (Ct <40) that were collected between October 8 and November 20, 2020 in Paris, before the detection of the B.1.1.7 variant in France. They were thus considered as being infected with non-B.1.1.7 strains, even if the samples were not sequenced. The second series corresponds to 227 samples (Ct<33) that were collected between January 7 and March 27, 2021. Seven additional samples with Ct>33 were also included. The second series includes 70 non-B.1.1.7 and 157 B.1.1.7 samples.

### Analysis of samples collected before B.1.1.7 spreading

We titrated viral infectivity in the first series of 200 patients. 107 patients (53%) harboured detectable viable virus in their nasopharyngeal swabs (Fig. 2a). The infectious titers were highly variable, ranging from 0 to >10^6^ IU/ml. To further characterize these variations, we plotted Ct values, days POS and levels of anti-SARS-CoV-2 IgG or IgA for each individual (Fig. 2a). Data were ranked from high to low infectious titers in RDT + and RDT-categories. The RDT-category was then ranked from low to high levels of SARS-CoV-2 IgG. There was a strong inverse correlation between infectious titers and viral RNA levels, defined by the Ct value (Fig. 2b). The median titer within positive individuals was 10^3^ IU/ml (Fig. 2c). The antigenic RDT was positive in 93% of samples with viable virus (Fig. 2a,c). We also identified a fraction of individuals without detectable viable virus that were RDT-positive (23%). This allowed us to estimate the sensitivity of 93% (95% CI: 86 to 96) and specificity of 78% (95% CI: 68 to 85) of the RDT to detect infectious samples in this cohort. As expected, samples positive in the infectivity assay displayed lower Ct values (median: 22) than negative samples (median 31.3) (Fig. 2d). This represents a 630-fold (=2^9.3^) increase in viral RNA levels associated with viral infectivity. The highest viral titers (>10^5^) corresponded to Ct values < 22. The median time of infectious virus shedding was 2 days (range -3 to 14 days, IQR 1 to 4 days). 67% of individuals displayed infectious virus 0 to 2 days POS. Specific antibodies started to be detected at 5-10 days POS in nasopharyngeal swabs, with some exceptions. There was a clear dichotomy between the detection of infectious virus and the presence of anti-SARS-CoV-2 IgG or IgA antibodies (Fig. 2a,b). This strongly suggests that the antibodies locally present in nasopharyngeal secretions can neutralize viral infectivity. These results further support previous reports indicating that infectious SARS-CoV-2 could only be isolated in individuals with low or undetectable neutralizing antibodies in their serum ^11,12^. We did not have access to serum samples in this cohort to compare levels of mucosal and circulating antibodies.

**Fig. 2.**
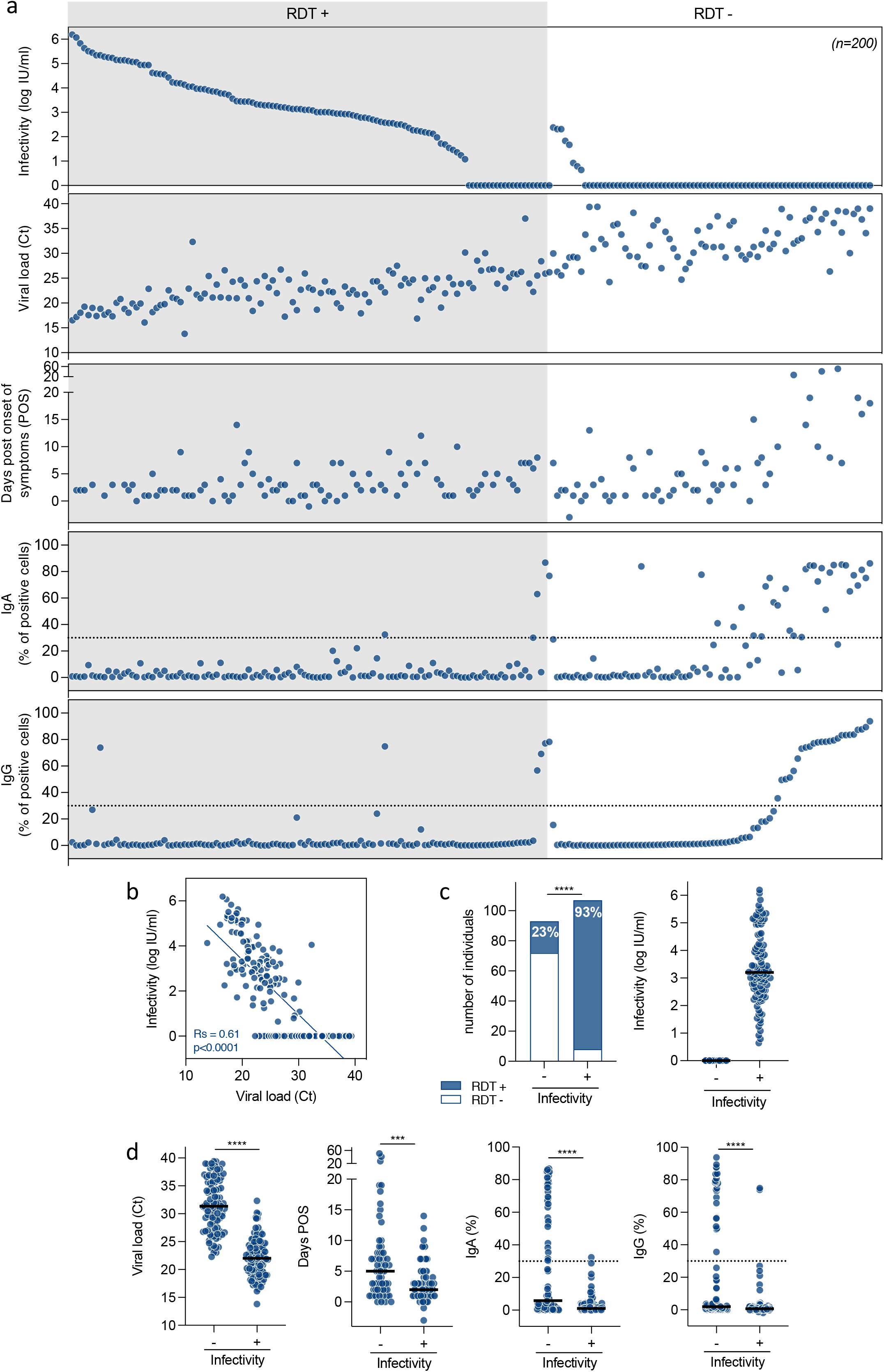
Analysis of 200 swabs collected from Oct to Nov 2020, before B.1.1.7 spreading. **a**. Infectious virus titers, viral RNA loads, days post onset of symptoms (POS), and anti-SARS-CoV-2 IgA and IgG levels are depicted in each panel. Each dot represents a sample. The samples were classified according to the RDT result (+ or -). The samples were then ranked from high to low infectivity. The samples without viable virus were ranked from low to high IgA or IgG levels. Dotted lines indicate the threshold for IgA or IgG positivity. **b**. Correlation between viral RNA loads (Ct) and infectivity (IU/ml). A Spearman correlation (SC) model was applied. SC p and r values are indicated. **c**. Analysis of RDT positivity and infectivity in the samples. Left panel: the number and % of samples with positive or negative RDT is indicated among non-infectious (-) or infectious (+) samples. Right panel: Infectious titers (IU/ml) among non-infectious (-) or infectious (+) samples. **d**. Viral RNA loads (Ct), days POS, levels of anti-SARS-CoV-2 IgA and IgG among non-infectious (-) or infectious (+) samples (from left to right panel). Dotted lines indicate the threshold for IgA or IgG positivity. A Mann-Whitney test was performed *** p<0.001, **** p<0.0001.

Altogether, these results highlight a strong variability in infectious shedding. The few individuals with the highest titres (>10^6^ IU/ml) may correspond to the rare super spreaders inferred by epidemiological surveys. These contact tracing surveys suggested that up to 90% of SARS-CoV-2 infections are spread by 10-20% of infected individuals ^22-24^.

### Comparative analysis of samples from non-B.1.1.7 and B.1.1.7 infected individuals

We then analyzed the second series of 70 non-B.1.1.7 and 157 B.1.1.7 patients (Fig. 3). The proportion of individuals positive with the RDT or displaying detectable infectious virus was similar with non-B.1.1.7 and B.1.1.7 variants (72 and 73% for RDT, and 42 and 39% for infectivity, respectively, Fig. 3a,b,d). 97% of individuals with viable virus were RDT positive (Fig. 3b). Median Ct values (21.3 and 20.6) were similar for the two viral subsets (Fig. 3c). As observed in the first cohort, there was a strong inverse correlation between infectious titers and Ct values for both variants (not shown). As expected, median Ct were lower in individuals carrying infectious virus, (19 and 17 for non-B.1.1.7 and B.1.1.7 variants, respectively, Fig. 3a,c). Intriguingly, among samples with viable virus, the median infectious titers were lower with B.1.1.7 compared to non-B.1.1.7 viruses (8.6 × 10^3^ and 0.4 × 10^3^ IU/ml, respectively) (Fig. 3d). The median time of infectious virus shedding was 2 days POS with both viruses (range -1 to 10 days) (Fig. 3e). Anti-SARS-CoV-2 IgG or IgA were not detected in nasopharyngeal swabs carrying viable virus, with a few exceptions (Fig. 3e). Overall, these results indicate that there is no significant difference between the two viral groups regarding the various parameters, when analyzed globally.

**Fig. 3.**
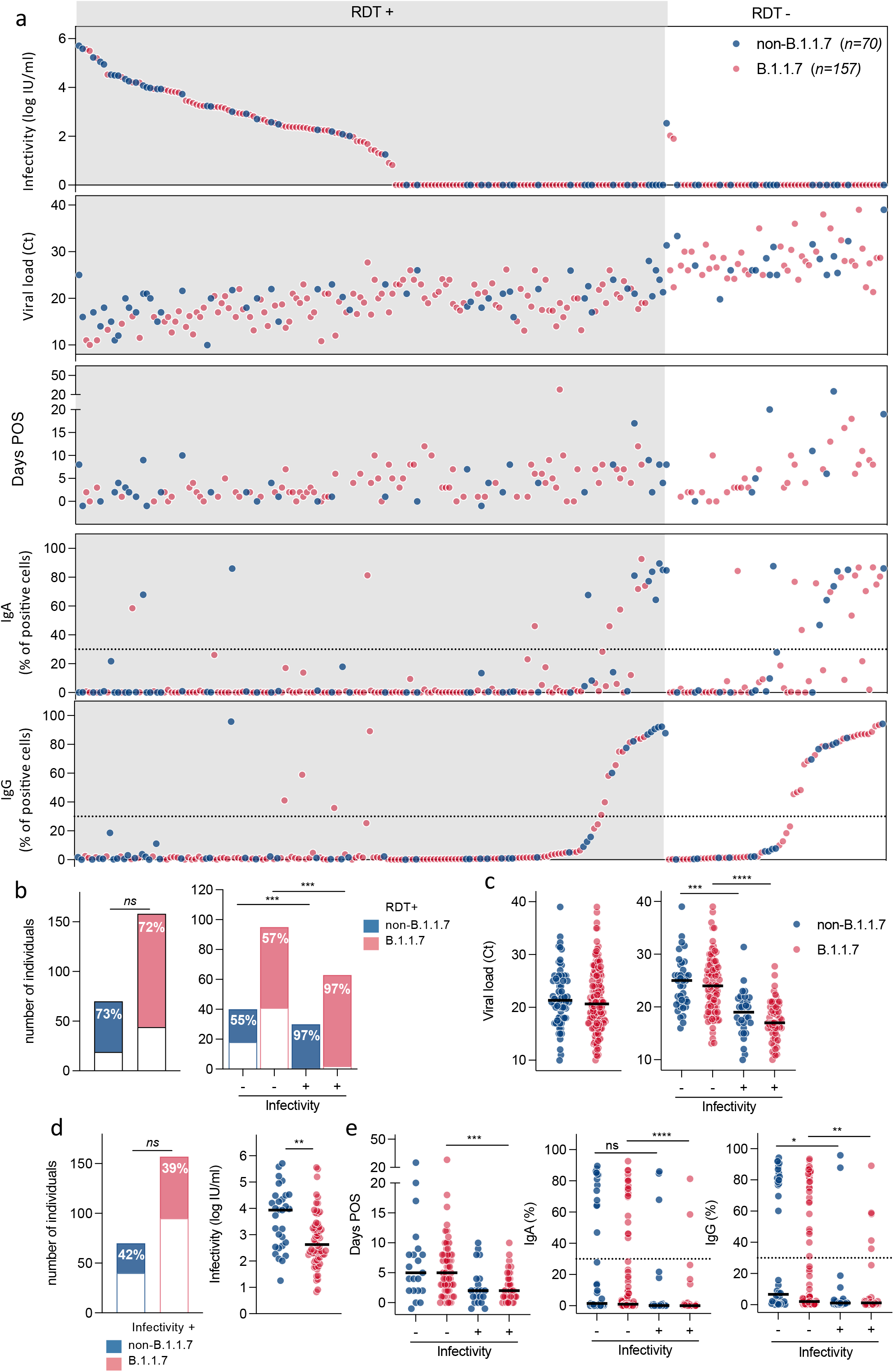
Comparison of nasopharyngeal swabs from 70 non-B.1.1.7 and 157 B.1.1.7-infected individuals. **a**. Infectious virus titers, viral RNA loads, days POS, and anti-SARS-CoV-2 IgA and IgG levels are depicted in each panel. Each dot represents a sample. Blue dots: non-B.1.1.7 viruses. Red dots: B.1.1.7 variant. The samples were classified according to the RDT result (+ or -). The samples were then ranked from high to low infectivity. The samples without viable virus were ranked from low to high IgG levels. Dotted lines indicate the threshold for IgA or IgG positivity. **b**. Proportion of individuals with a RDT positive test. Left panel: Proportion of individuals with a RDT positive test. Right panel: Proportion of individuals with a RDT positive test among those carrying (+) or not carrying (-) infectious virus. A Fisher’s test was performed. **c**. Viral RNA loads (Ct) in all samples (left panel) or among non-infectious (-) and infectious (+) samples (left and right panels, respectively). **d**. Analysis of infectivity. Left panel: the number and % of samples with infectious virus is indicated. Right panel: Infectious titers (IU/ml) among samples harboring infectious (+) virus. **e**. Days POS, levels of anti-SARS-CoV-2 IgA and IgG among non-infectious (-) or infectious (+) samples (from left to right panel). Dotted lines indicate the threshold for IgA and IgG positivity. A Mann-Whitney test was performed to compare non-B.1.1.7 and B.1.1.7 samples (c, left panel, d, right panel). A Kruskal-Wallis was performed to compare non-B.1.1.7 and B.1.1.7 in infectious versus non-infectious samples. ns: non-significant, **: p<0.01, ***: p<0.001, **** p<0.0001.

We then evaluated by a multivariate analysis (partial least square regression) the associations between infectious titers and other parameters (Fig. 4a). The characteristics associated with detection of viable virus were RDT positivity, low Ct values, early time since onset of symptoms, absence of IgG or IgA antibodies. There was no association between detection of viable virus and disease severity, age or sex of the individuals (Fig. 4a). The same profile of associations was observed in individuals infected with non-B.1.1.7 and B.1.1.7 variants (Fig. 4a and data not shown).

**Fig. 4.**
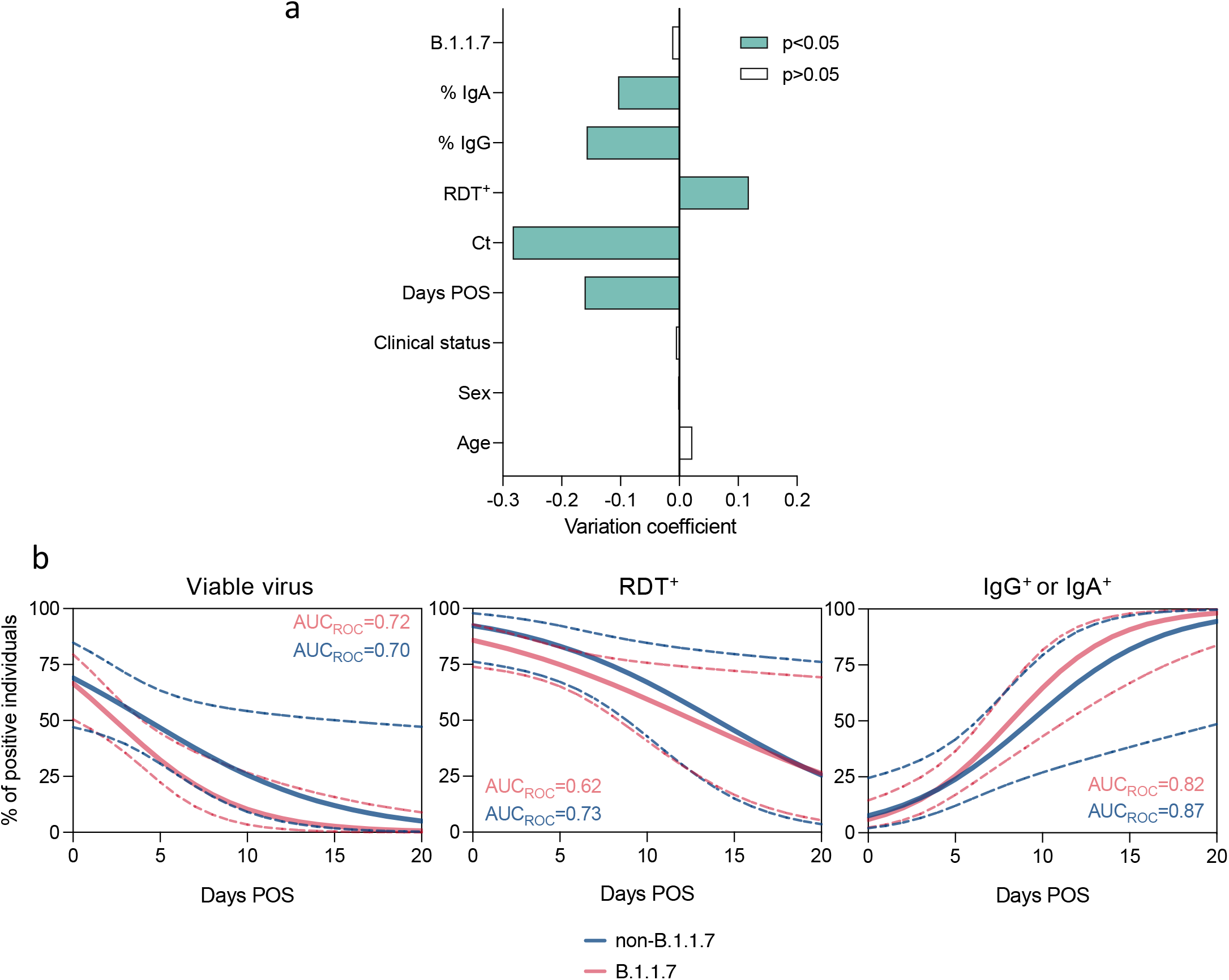
Associations between infectious viral titers and other parameters. The analysis was performed on swabs from 70 non-B.1.1.7 and 157 B.1.1.7-infected individuals. **a**. Parameters associated with viral titer analyzed using the partial least squares (PLS) regression method. The variation coefficient is the estimate effect of each standardized parameter on the standardized infectious titer. Negative and positive coefficients denote negative and positive associations, respectively. Significant associations are in green. Significance of the variation coefficients was assessed with Jackknife approximate t tests. **b**. Logistic regressions of the percentage of positive individuals for each test, depending on days POS. Models are in blue for non-B.1.1.7 and in red for B.1.1.7. The 95% confidence intervals are represented by dashed lines. The AUCROC indicates the goodness of fit, the closest to 1 the better the prediction is. Curves were compared using a Chi-square test of regression with time and variant parameters.

### Duration and extent of viral shedding in nasopharyngeal swabs from non-B.1.1.7 and B.1.1.7 infected individuals

Viral shedding may begin 5 to 6 days before the appearance of the first symptoms ^25-27^. After symptom onset, viral loads decrease monotonically ^25-27^. We measured the proportion of individuals with viable virus, at each time point, from 0 to 20 DOS. We performed this analysis on the second cohort, to compare non-B.1.1.7 and B.1.1.7 infected individuals. Fitted logistic regression distribution curves indicated that the proportion of individuals with viable virus was roughly similar with B.1.1.7 and non-B.1.1.7 (Fig. 4b). However, the slope of decrease appeared steeper with B.1.1.7 samples (although not significantly different from non-B.1.1.7 samples) suggesting that the peak of infectivity may have occurred earlier, before onset of symptoms. The absence of B.1.1.7 samples collected at the pre-symptomatic phase precluded this analysis. The evolution of RDT positivity and appearance of SARS-CoV-2 antibodies overtime was similar for B.1.1.7 and non-B.1.17 (Fig. 4b).

These results indicate no major difference regarding the evolution of the proportion of individuals with viable virus in this cohort. The steeper decrease of detection of viable virus in B.1.1.7-infected individuals may be linked to the slightly reduced viral titers observed with this variant in the whole cohort (Fig. 3c).

### Nasopharyngeal cytokine responses in non-B.1.1.7 and B.1.1.7 infected individuals

Perturbed systemic production of cytokines is a hallmark of disease severity in COVD-19 patients ^28-31^. It has also been reported a dysbiosis and a modification of cytokine profile at mucosal surfaces in severe patients ^18^. In this previous study, performed on samples from individuals infected before B.1.1.7 spreading, cytokine responses were compartmentalized ^18^. At least 13 nasopharyngeal cytokines (VEGF, FGF, IL-1RA, IL-6, TNFα, IL-10, CCL2/MCP-1, CXCL10/IP-10, CCL3/MIP-1, CCL19/MIP-3, PD-L1, G-CSF and Granzyme B) were differently regulated between critical and non-critical patients^18^. To better understand the host response to the different variants, we measured the levels of 46 cytokines in nasopharyngeal specimen from individuals infected with either non-B.1.1.7 and B.1.1.7 viruses. A set of 17 cytokines (IFN-β, IP-10, CD40L, PGDF-AB, GM-CSF, IL-3, eotaxin, RANTES, Granzyme B, IL-12p70, IL-4, IL-5, IL-13, IL-1RA, IL-10, PD-L1) was significantly increased in B.1.1.7 samples. Examples of increased cytokines, including IFN-β, IP-10, IL-10, and TRAIL are depicted in Fig. 5a. Other cytokines, such as IFNα2, IL-6, IL-17a and TNFα were non significantly elevated (Fig. 5a). The full set of results appear in Fig. S1. Moreover, there was a strong negative correlation between Ct values and levels of a few cytokines, such as IFNα2 (Fig. 5b). This correlation was observed with both non-B.1.1.7 and B.1.1.7 variants. This was not the case for other cytokines, such as IL-6 (Fig. 5b).

**Fig. 5.**
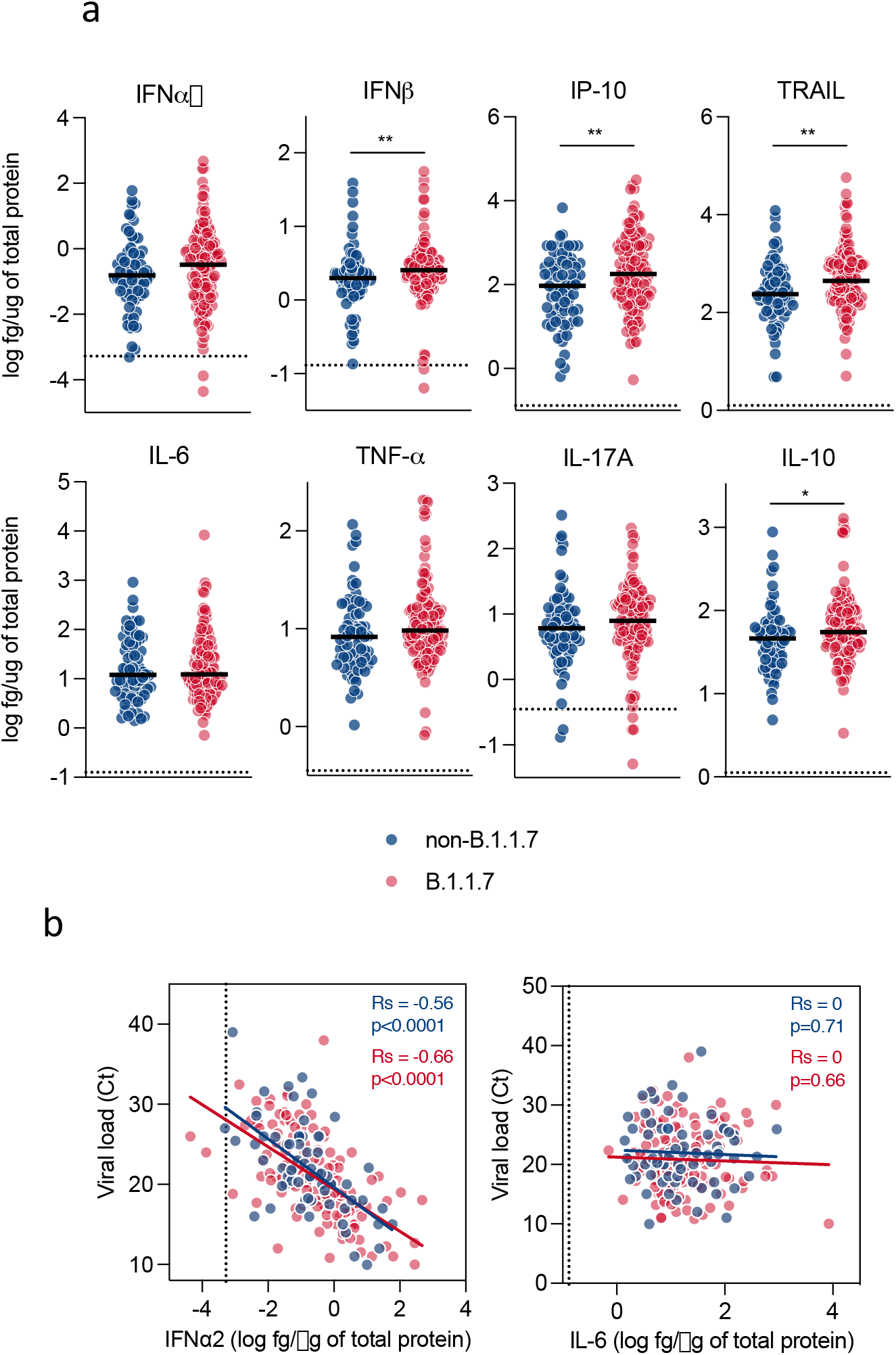
Cytokines production in swabs from non-B.1.1.7 and B.1.1.7 infected individuals. A panel of 46 cytokines produced in nasopharyngeal swabs from 70 non-B.1.1.7 and 132 B.1.1.7-infected individuals was measured using a bead-based multiplex immunoassay system Luminex or a digital Elisa Simoa (IFNα2, IFNλ, IFNλ3, IL-17A). **a**. Comparison of the levels of 8 cytokines. The full set of results is depicted in Fig. S1. Each dot represents a sample. Blue dots: non-B.1.1.7 viruses. Red dots: B.1.1.7 variant. Dotted lines indicate the limits of detection. p values were determined with the Mann-Whitney test between non-B.1.1.7 and B.1.1.7 cases. *p < 0.05; **p < 0.01; ***p < 0.001. **b**. Correlations between viral RNA loads (Ct) and concentrations of IFNα2 (left panel) and IL-6 (right panel). The Spearman correlation (SC) model was applied for non-B.1.1.7 (blue dots) and B.1.1.7 (red dots) samples. SC p and r values are indicated.

These results strongly suggest that B.1.1.7 infection may cause higher inflammatory and interferon responses.

### Combined analysis of immunological and virological parameters

We then established a two-by-two correlation table of all virological and immunological parameters that were measured in the second cohort, independently of the viral variant (Fig. S2). Infectious viral titers inversely correlated with Ct values, days POS and anti-SARS-CoV-2 antibodies. Ct values negatively correlated with the levels of IFNα2, as depicted in Fig. 5a, and with GRO-B, but not with any other cytokines. There were strong correlations between most of the up-regulated cytokines, suggesting that similar pathways of activation drove their production (Fig. S2).

## Discussion

We measured infectious SARS-CoV-2 in nasopharyngeal swabs with a novel cell reporter system termed S-Fuse assay, that we improved by adding TMPRSS2 ^19,20^. The technique is based on the detection of GFP+ syncytia formed between infected and neighbouring cells. The assay is rapid, and infectivity is revealed in less than 24h. It is semi-automated and less labour intensive than classical assays based on Vero cells. It can be applied to various types of clinical samples: our preliminary experiments indicate for instance that viable virus is detectable in bronchoalveolar lavages from COVID-19 hospitalized patients (not shown). We combined the S-Fuse assay with RT-qPCR, lateral flow antigenic RDT, measurement of anti-SARS-CoV-2 antibodies and cytokines, in order to provide a global overview of the duration and determinants of infectious viral shedding in 427 COVID-19 patients. Overall, 197/427 (46%) of the participants displayed infectious virus. Kampen et al recently reported the isolation of SARS-CoV-2 in 62/690 samples (9%) from COVID-19 hospitalized patients, by using Vero cells ^11^. This strongly suggests that the S-Fuse assay is particularly sensitive to detect viable virus.

We report a strong correlation between detection of viable virus, and RDT positivity or low Ct values. About 95% of samples carrying infectious virus were RDT+ and displayed Ct values <22. Therefore, these two parameters are reliable markers of the presence of infectious virus, at least before the appearance of SARS-CoV-2 antibodies. Despite evidence of prolonged viral RNA shedding in some specimen (up to 28 days), viable virus detection was rather short lived in our study. The low amounts of viral RNA released on the long-term thus likely corresponds to viral remnants rather than to infectious virus. The samples carried viable virus mostly within the first 10 days, with a median of 2-3 days POS and occasional detection up to 14 days. In other reports, no live virus was detected in respiratory samples after 8-9 days of symptoms, probably reflecting the lower sensitivity of Vero cell-based assays ^7-9,13,32,33^. Of note, shedding of infectious virus up to 20 days in one severe COVID-19 case and 70 days in one immunocompromised patient has also been reported ^6,34^.

The extended detection of viable virus reported here suggests that the contagiousness period can be slightly longer than previously thought, at least for some individuals. This may have important implications for viral transmission in a community or hospital setting. Moreover, our results confirm that the antigenic test has a better positive predictive value than a RT-qPCR test for the potential contagiousness of a given individual ^15^.

We then compared the virological and immunological characteristics of nasopharyngeal swabs from symptomatic individuals infected with non-B.1.1.7 and B.1.1.7 variants. There was a non-significant trend to a reduction of Ct in infectious samples carrying B.1.1.7, when compared to non-B.1.1.7 viruses. The proportion of individuals displaying positive antigenic tests was similar with non-B.1.1.7 and B.1.1.7. The range of infectious viral titers were roughly similar with the different variants. Unexpectedly, the observed trend was a decrease, rather than an increase of viral titers in samples from B.1.1.7-infected individuals. The decrease overtime of the proportion of individuals carrying infectious virus was also somewhat steeper upon B.1.1.7 infection. The reasons for this remain unclear. This may suggest that infectivity peaked earlier, before onset of symptoms. It is also possible that some cytokines modulate infectivity of the samples. An analysis performed on a larger number of symptomatic and asymptomatic individuals will help determining whether B.1.1.7’s increased transmissibility is associated with extended or higher shedding of viral RNA after onset of symptoms, as reported in recent studies ^3-5,35^. The differences between viral lineages were however minimal in some of these studies. Future work is also warranted to assess viral loads in asymptomatic B.1.1.7-infected individuals.

We report here a main difference between B.1.1.7 and other lineages in the nasopharyngeal swabs, a higher release of 17 cytokines out of a panel of 46 measured cytokines. This raises interesting questions about the origin and consequences of this phenomenon. Higher or disequilibrated cytokine responses were associated with disease severity, in studies performed before the spreading of B.1.1.7 variant ^28-31^. It remains yet unclear whether B.1.1.7 is more pathogenic than pre-existing viruses. It has been initially reported an increased mortality in community-tested cases of B.1.1.7 ^1^ but other studies did not indicate a clear clinical effect associated with B.1.1.7 infection ^2 5^. In the hamster model, B.1.1.7 infection did not cause more severe disease or higher viral loads than previously circulating strains ^36^ 37. However, some cytokines (including IL-6, IL-10 and IFN-γ) were most pronouncedly up-regulated in B.1.1.7 infected hamsters as compared to three other strains ^37^. Our observations are in line with the results obtained in this animal model. Whether elevated levels of cytokines at the nasopharyngeal mucosae may influence viral transmissibility or reflects differences in the kinetic or extent of viral shedding will deserve further exploration. It will be also worth determining whether cytokines are elevated in blood samples from individuals infected with B.1.1.7, since a tissue compartmentalization of SARS-CoV-2 immune responses have been recently reported, with a role for the nasopharyngeal microbiome in regulating local and systemic immunity that determines COVID-19 clinical outcomes ^18^.

Our study has some limitations. We did not analyse longitudinal samples and the number of samples collected at late time points (after day 10) was scarce. We also did not assess systemic immune responses, since we did not have access to the blood samples of the same individuals. It will be worth determining whether the oral cavity and saliva, important sites for SARS-CoV-2 transmission^38^, carry different levels of variants. Future work with higher numbers and types of specimen and serial sampling will help further characterizing the virological and immunological determinants of viral transmission. It will be also of great interest to analyze these parameters in asymptomatic and pre-symptomatic individuals, who play a prominent role in viral spreading at the population level.

## Data Availability

All data supporting the findings of this study are available within the paper and are available from the corresponding author upon request. The viral sequences have been deposited to GISAID and accession numbers are available upon request.

## Methods

No statistical methods were used to predetermine sample size. The experiments were not randomized and the investigators were not blinded to allocation during experiments and outcome assessment. Our research complies with all relevant ethical regulation.

### Hôpital Européen Georges Pompidou (HEGP) cohorts

The first cohort is composed of SARS-CoV-2 positive samples for which PCR was performed at the HEGP Virology Laboratory with the AllplexTM 2019-nCoV Assay - Seegene Inc between October 8 and November 18, 2020. The Ct analysis was performed for the N gene. The second cohort (n=227) includes all positive samples from HEGP between January 7 and March 21, 2021 that were compatible with variant screening (Ct<33) according to national recommendations. 210 Samples were subjected to specific molecular screening for N501Y, E484K, K417N and V1176F mutations (VirSNiP assay, Tib Molbiol, Berlin, Germany). 17 samples were classified as B.1.1.7 based on the absence of S gene amplification by TaqPath™ COVID 19 RT PCR Kit (Thermo Fisher Scientific, Coutaboeuf, France). Full-length viral genomes were obtained on 183 samples (59 non-B1.1.7 and 124 B.1.1.7) out of 227,by Illumina sequencing (Artic protocol, NEB library) and confirmed the PCR analysis. The N gene result was used for Ct analysis (AllplexTM SARS-CoV-2 Assay - Seegene Inc). The transport medium was the Yocon Virus Sampling kit (Yocon Biology Company, Beijing, China). Demographic information was collected (sex, age). The severity of the disease at the time of sampling was classified as asymptomatic, mild (symptoms without hospitalization), moderate (hospitalization), critical (intensive care). The date of onset of symptoms was collected when available. Of note, we did not mix the Ct values obtained in the two cohorts, since different PCR tests were used.

### Ethics statement

Our observational work was carried out in accordance with the Declaration of Helsinki with no sampling addition to usual procedures. Swab specimens were obtained for standard diagnostic following medical prescriptions in our hospital. The project was evaluated by the ethics committee “Comité d’éthique de la recherche AP-HP Centre“ affiliated to the AP-HP (Assitance publique des Hopitaux de Paris). It obtained an approval (IRB registration # 00011928) on February 17, 2021.

### Lateral Flow Antigenic Rapid Diagnostic Rapid Test (RDT)

COVID-19 Ag test (SD Biosensor, Inc, Republic of Korea) was used on transport media according to the manufacturer’s recommendations.

### S-Fuse assay

U2OS-ACE2 GFP1-10 or GFP 11 cells, also termed S-Fuse cells, become GFP+ when they are productively infected by SARS-CoV-2 ^19,20^. TMPRSS2 was added in S-Fuse cells as described, yielding S-Fuse-T cells ^19^. Cells were tested negative for mycoplasma. Cells were mixed (ratio 1:1) and plated at 8×10^3^ per well in a μClear 96-well plate (Greiner Bio-One). The nasopharyngeal swabs or a SARS-CoV-2 strain control were added to the S-fuse cells at serial dilutions from 1:10 to 1:1 000 000. 18 hours later, cells were fixed with 2% PFA, washed and stained with Hoechst (dilution 1:1,000, Invitrogen). Images were acquired with an Opera Phenix high content confocal microscope (PerkinElmer). The GFP area and the number of nuclei were quantified using the Harmony software (PerkinElmer). The viral titer (Infectious units /mL) was calculated from the last positive dilution with 1 infectious unit (IU) being 3 times the background (GFP area in non-infected controls).

### S-Flow Assay

The S-Flow assay was performed as described ^17,39^. Briefly, HEK-293T (referred as 293T) cells were acquired from ATCC (ATCC® CRL-3216TM) and tested negative for mycoplasma. 293T Cells stably expressing Spike or control cells were transferred into U-bottom 96-well plates (10^5^ cells/well). Cell were incubated at 4°C for 30 min with nasopharyngeal swabs (1:5 dilution) in PBS containing 0.5% BSA and 2 mM EDTA, washed with PBS, and stained using anti-IgG AF647 or Anti-IgA AF647 (Jackson ImmunoResearch). Cells were washed with PBS and fixed 10 min using 4% PFA. Data were acquired on an Attune Nxt instrument (Life Technologies). Stainings were also performed on control (293T Empty) cells. Results were analyzed with FlowJo 10.7.1 (Becton Dickinson). The positivity of a sample was defined as a specific binding above 20%. The specific binding was calculated as follow: 100 x (% binding 293T Spike - % binding 293T Empty)/ (100 - % binding 293T Empty).

### Anti-SARS-CoV-2 antibody and Virus stock

The human anti-SARS-CoV2 monoclonal antibody mAb48 recognizes the RBD ^20^. SARS-CoV-2 isolate (Wuhan) was used as a positive control as described ^19 20^.

### Measurement of cytokines

Nasopharyngeal samples were inactivated with Triton X-100 1% (v/v) for 2h at RT, and cytokines were quantified as described ^18^. IFNα2, IFNγ, IFNλ3, IL-17A were quantified with Simoa assays developed as described with Quanterix Homebrew kits ^40^. Other cytokines and chemokines were measured with a commercial Luminex multi-analyte assay (Biotechne, R&D system).

### Statistical analysis

Flow cytometry data were analyzed with FlowJo v10 software (TriStar). Calculations were performed using Excel 365 (Microsoft). Figures were drawn on Prism 9 (GraphPad Software). Statistical analysis was conducted using GraphPad Prism 9. Statistical significance between different groups was calculated using the tests indicated in each figure legend.

Partial least square regression with “leave-one-out” strategy was performed with R version 4.0.2 on RStudio Desktop 1.3.959 (R Studio, PBC) using the Package pls version 2.7-3. Briefly, numerical data were centered and scaled before performing the regression and the number of components was selected with the *selectNcomp* function. Significance of the variation coefficients was assessed with Jackknife approximate t tests (*jack*.*test*). We performed logistic regressions on different parameters depending on time since onset of symptoms with the *glm* function of the stats package. The goodness of fit was assessed by calculating the area under the ROC curve between observed and predicted results (AUCROC) (package pROC: http://expasy.org/tools/pROC). We compared curves using a Chi-square test of the regression with time and variant parameters. For other R analysis we used the following packages: corrplot (https://github.com/taiyun/corrplot) and readxl (https://CRAN.R-project.org/package=readxl)

## Acknowledgments

We thank Michael Maaran Rajah, Nicoletta Casartelli, Michael White and Jeremy Dufloo for discussion and critical reading of the manuscript. We thank AGEPS, Assistance Publique - Hôpitaux de Paris, for providing the RDTs. We thank the patients who participated in the study, the clinical staff involved in their management and Théo Hirsch for the stimulating discussions. We thank the members of the Virus and Immunity Unit for discussion and help, Nathalie Aulner and the UtechS Photonic BioImaging (UPBI) core facility (Institut Pasteur), a member of the France BioImaging network, for image acquisition and analysis, and Cyril Planchais & Hugo Mouquet for the kind gift of mAb48.

## Author contributions

Experimental strategy design, experiments: BM, DP, LG, NS, NR, IS, PG, FP, FGB, TB, HP, DV, OS

Provide access to biological samples and clinical information: NDG, JR, JP, VE, LB, GO, LN, VM, JF, MW, SI, HP, DV

Manuscript writing: DP, LG, TB, OS

Manuscript editing: BM, NS, JPDS, HP, DV

## Figure legends

**Fig. S1 (related to Fig. 5). Cytokines production in swabs from non-B**.**1**.**1**.**7 and B**.**1**.**1**.**7 infected individuals**.

A panel of 46 cytokines produced in nasopharyngeal swabs from 70 non-B.1.1.7 and 132 B.1.1.7-infected individuals was measured using a bead-based multiplex immunoassay system Luminex or the digital Elisa Simoa (IFNα2, IFNγ, IFNλ3, IL-17A). Dotted lines indicate the limits of detection. p values were determined with the Mann-Whitney test between non-B.1.1.7 and B.1.1.7 cases. *p < 0.05; **p < 0.01; ***p < 0.001.

**Fig. S2. Pearson correlation matrix of features assessed in individuals infected with non-B**.**1**.**1**.**7 or B**.**1**.**1**.**7 viruses**. Features were ordered using hierarchical clustering. Only statistically significant correlations (p<0.05) are depicted. Positive correlations are indicated in red and negative correlations in blue. Color intensity and size of the dots are proportional to the p value.

**Table S1**. Characteristics of the individuals with PCR-confirmed SARS-CoV-2 infection.

Characteristics of the 200 RT-qPCR diagnosed COVID-19 individuals collected between October 8 and November 20, 2020 (upper panel) and 227 individuals collected between January 7 and March 27, 2021 (70 infected with non-B.1.1.7 and 157 infected with B.1.1.7) (lower panel).

