## Supplementary Figures for "Release of infectious virus and cytokines in nasopharyngeal swabs from individuals infected with non-B.1.1.7 or B.1.1.7 SARS-CoV-2 variants"

| Cohort #1 | | |
|---|---|---|
| Number of samples | | 200 |
| Age (years), median (IQR) | | 47 (29, 67) |
| Sex | | |
| | Female | 114 |
| | Male | 83 |
| | ND | 3 |
| Clinical | | |
| | Asymptomatics | 18 |
| | Mild symptoms | 114 |
| | Hospitalized | 34 |
| | ICU | 11 |
| | ND | 23 |
| Number of samples with known days POS | | 149 |
| Days POS, median (IQR) | | 3 (1, 6) |

| Cohort #2 | | Non-B.1.1.7 | B.1.1.7 |
|---|---|---|---|
| Number of samples | | 70 | 157 |
| Age (years), median (IQR) | | 66 (48, 85) | 55 (40,74) |
| Sex | | | |
| | Female | 42 | 95 |
| | Male | 28 | 62 |
| Clinical | | | |
| | Asymptomatics | 5 | 10 |
| | Mild symptoms | 21 | 52 |
| | Hospitalized | 19 | 43 |
| | ICU | 11 | 26 |
| | ND | 14 | 26 |
| Number of samples with known days POS | | 43 | 108 |
| Days POS, median (IQR) | | 3 (1,8) | 3 (1.8, 7) |

Supplemental Table 1

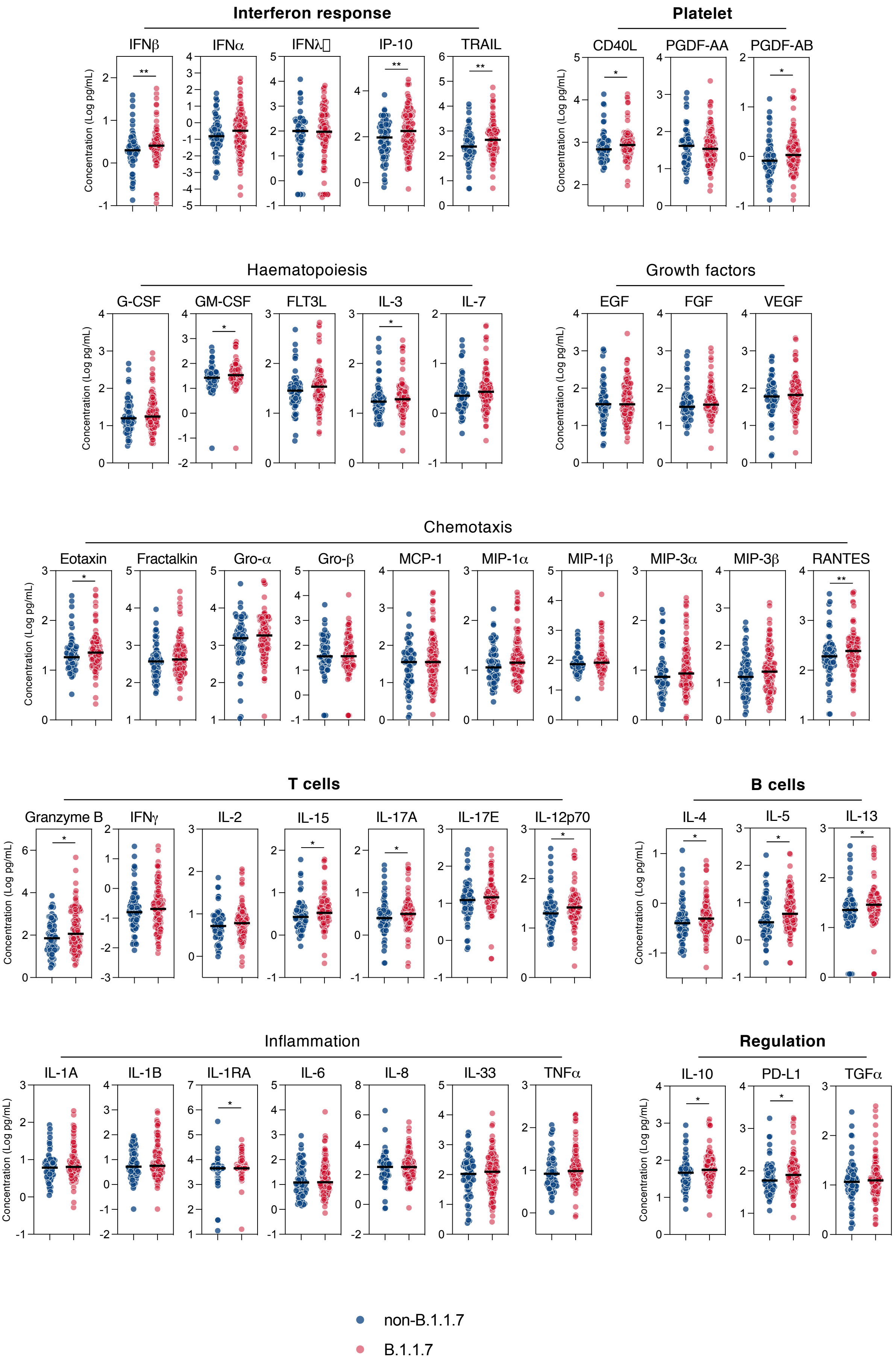
Interferon response
IFNβ
IFNα
IFNλ
IP-10
TRAIL
Platelet
CD40L
PGDF-AA
PGDF-AB
Haematopoiesis
G-CSF
GM-CSF
FLT3L
IL-3
IL-7
Growth factors
EGF
FGF
VEGF
Chemotaxis
Eotaxin
Fractalkin
Gro-α
Gro-β
MCP-1
MIP-1α
MIP-1β
MIP-3α
MIP-3β
RANTES
T cells
Granzyme B
IFNγ
IL-2
IL-15
IL-17A
IL-17E
IL-12p70
B cells
IL-4
IL-5
IL-13
Inflammation
IL-1A
IL-1B
IL-1RA
IL-6
IL-8
IL-33
TNFα
Regulation
IL-10
PD-L1
TGFα
Concentration (Log pg/mL)
non-B.1.1.7
B.1.1.7

Figure S1

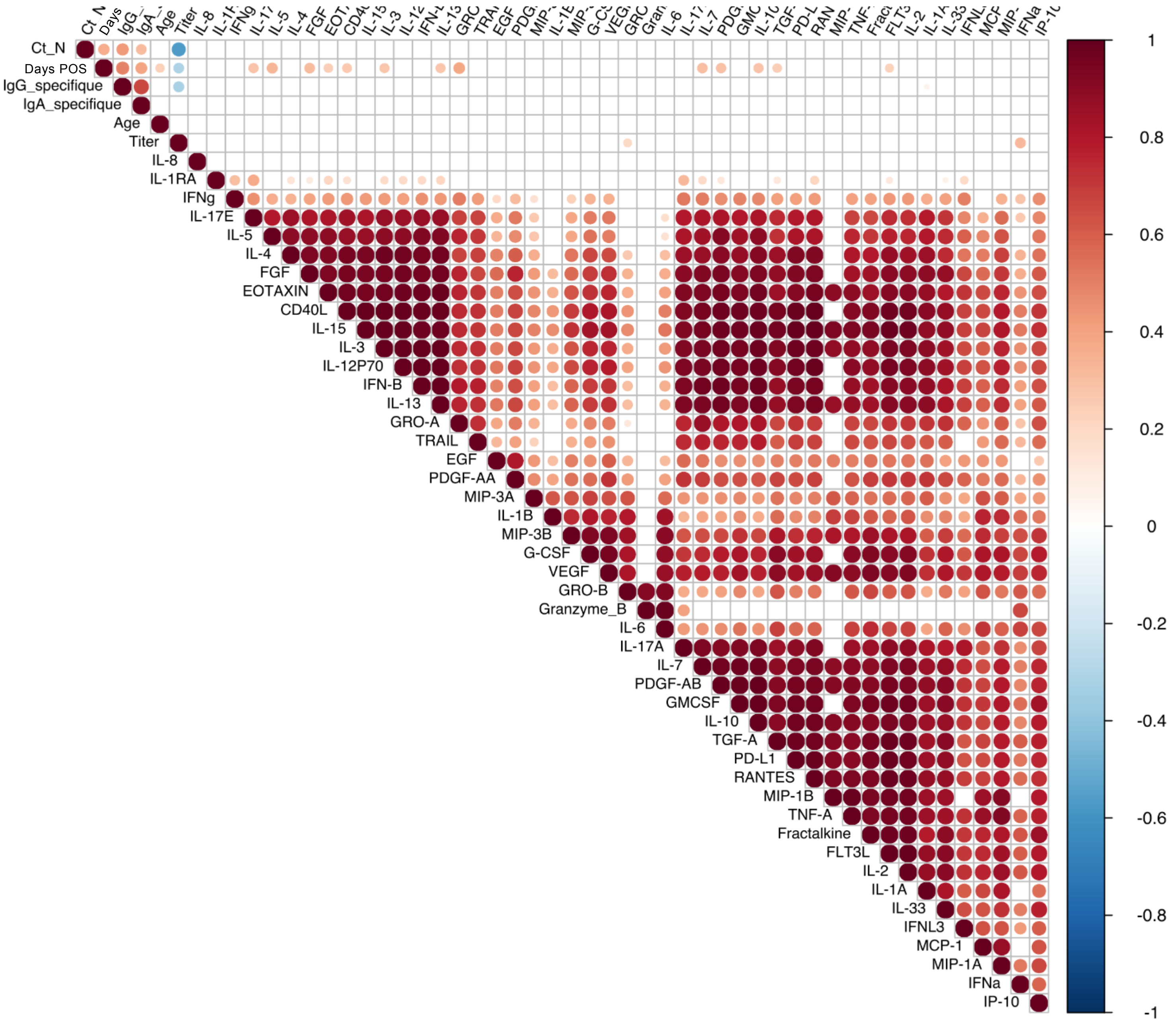
Ct_N
Days POS
IgG_specifique
IgA_specifique
Age
Titer
IL-8
IL-1RA
IFNg
IL-17E
IL-5
IL-4
FGF
EOTAXIN
CD40L
IL-15
IL-3
IL-12P70
IFN-B
IL-13
GRO-A
TRAIL
EGF
PDGF-AA
MIP-3A
IL-1B
MIP-3B
G-CSF
VEGF
GRO-B
Granzyme_B
IL-6
IL-17A
IL-7
PDGF-AB
GMCSF
IL-10
TGF-A
PD-L1
RANTES
MIP-1B
TNF-A
Fractalkine
FLT3L
IL-2
IL-1A
IL-33
IFNL3
MCP-1
MIP-1A
IFNa
IP-10
1
0.8
0.6
0.4
0.2
0
-0.2
-0.4
-0.6
-0.8
-1

Figure S2
